# Early Diagnosis and Clinical Significance of Acute Cardiac Injury - Under the Iceberg: A Retrospective Cohort Study of 619 Non-critically Ill Hospitalized COVID-19 Pneumonia Patients

**DOI:** 10.1101/2020.07.06.20147256

**Authors:** Yang Xie, Sichun Chen, Xueli Wang, Baige Li, Tianlu Zhang, Xingwei He, Ningling Sun, Luyan Wang, Hesong Zeng, Yin Shen

**Affiliations:** Department of Cardiology, Tongji Hospital, Tongji Medical college, Huazhong University of Science and Technology, Wuhan, China; Medical Research Institute, Wuhan University Renmin Hospital, Wuhan University, Wuhan, China; Institute of Central China Development, Wuhan University, Wuhan, China; Heart Center, Peking University Peoples Hospital, Beijing, China

**Keywords:** COVID-19, Acute cardiac related injury, hypertension, PLT, NLR

## Abstract

**Rationale:** Coronavirus disease 2019 (COVID-19) can cause a viral pneumonia together with other extrapulmonary complications. Acute cardiac related injury (ACRI) is common in hospitalized COVID-19 patients.

**Objective:** To explain the pathological mechanism of ACRI and improve the treatment strategy by retrospectively observing the factors associated with ACRI and factors affecting the prognosis of ACRI with COVID-19 at an early stage.

**Methods:** 619 COVID-19 patients were from Tongji Hospital, Wuhan. Student’s t test was used for continuous variables while Pearson χ^2^ test for categorical factors. Univariable and multivariable logistic regression models were applied to estimate odds ratio (OR) with 95% confidence interval (CI).

**Results:** Among the 619 OOS Level-I hospitalized COVID-19 patients, 102 (16.5%) were defined as ACRI (stage-1: 59 cases, stage-2: 43 cases). 50% of ACRI patients developed into severe cases and 25 patients died(CFR=24.5%), 42 times that of non-ACRI patients. Elderly (OR=2.83, *P*<0.001), HTN (OR=2.09, *P*=0.005), γ-globulin (OR=2.08, *P*=0.004), TCM (OR=0.55, *P*=0.017), PLT (OR=2.94, *P*<0.001) and NLR (OR=2.20, *P*=0.004) were independently correlated with ACRI. SBP ⩾ 140, dyspnea, DM, smoking history were correlated with ACRI-stage2 only. In the prognostic subgroup analysis of ACRI patients, γ-globulin treatment could prolong LOS (29.0 ± 7.2 days Vs 23.5 ± 8.1 days, *P*=0.004). TCM (OR=0.26, *P*=0.006), SBP ⩾ 160 (OR= 22.70, *P*=0.005), male (OR=2.66, *P*=0.044) were associated with severe illness while corticosteroids treatment (OR=3.34, *P*=0.033) and male (OR=4.303, *P*=0.008) with death. Surprisingly, we found the mortality of non-elderly patients is higher than elderly (32.4% VS 20.0%, *P*=0.164), and both IKF and RASI treatment were not correlated with any prognostic indicators including severe, death and LOS.

**Conclusion:** This study observed that several non-traditional issues were associated with early cardiac injury in COVID-19 while many traditional cardiovascular risk factors were not. Besides elderly and male, hypertension was confirmed to be the most important risk factor.

## Background

Coronavirus disease 2019 (COVID-19) has infected >8.0 million people worldwide and killed >450 000 as of June 22 according to the latest statistics from Johns Hopkins University. Although most patients died from acute respiratory failure, several studies discovered that acute cardiac injury, which would process to heart failure, is common in COVID-19 patients^1^. Several studies have reported that acute cardiac injury (ACI) might occur after respiratory viruses infection such as SARS-CoV^1^, influenza^2^, MERS-CoV^3^. Acute myocardial injury (AMI) and some of its complications were also observed in 9.5% of all COVID-19 patients in Italy (up to April 2020) according to NICE guidline^*4*^. In a recent single-center study of 416 patients with confirmed COVID-19, cardiac troponin I (cTnI) elevation was seen in 19.7% patients and corresponded to higher in-hospital mortality^5^, indicating that AMI may affect the prognosis of COVID-19 patients. A meta-analysis about the prognostic value of several cardiac biomarkers in COVID-19 patients discovered that patients who died or were seriously ill had higher cTnI and CK levels than those who survived or were not critically ill ^6^. One study from Renmin Hospital of Wuhan University showed among 671 hospitalized patients, a total of 62 patients (9.2%) died, who had myocardial injury more often (75.8% vs 9.7%, *P* < 0.001) than survivors^7^. A retrospective case series analysis demonstrated that myocardial injury is significantly associated with fatal outcome of COVID-19. In total, 187 patients with confirmed COVID-19 were involved in the research and those with cardiovascular disease (CVD) and normal TnT levels experienced a more promising prognosis compared with those who had escalated TnT levels but without CVD (mortality, 13.3% vs 37.5%), which indicated that myocardial injury played a more important role in the fatal outcome of COVID-19 than CVD^8^.

Although scientists haven’t explained the exact pathophysiological mechanism of ACI by COVID-19, since the two viruses are highly homologous in genome, COVID-19 might share the mechanism with SARS-COV. In a previous research, the genome of SARS-CoV was detected in the heart of 35% patients infected, which increased the possibility that SARS-CoV could damage myocardial cell directly^9^. In a recent study, there was a significant linear positive correlation between plasma TnT level and plasma hypersensitive C-reactive protein level, suggesting that myocardial injury may be closely related to the pathogenesis of inflammation in disease progression. The virus particles spread through the mucous membrane of the respiratory tract and infect other cells, triggering cytokine storms and a series of immune responses. Huang et al. have emphasized that in COVID-19 patients, an imbalance in Th1 and Th2 responses leads to cytokine storms, which results in ACI. The theory of cytokine storms can be supported by increased level of d-dimer, IL-6, CRP, and LDH^10^. After inflammatory cytokines enter the blood, coronary blood flow and oxygen supply will decrease. Some scientists are curious about the role inflammatory markers play in ACI and hope that this could be a potential target for treatment. Besides, the gradual building up of unstable coronary plaque and microthrombus might contribute to ACI as well^11^.

Angiotensin-converting enzyme 2 (ACE2) is a membrane-bound aminopeptidase that is highly expressed in the heart and lungs and is critical in both cardiovascular and immune systems. More importantly, it has been identified as a functional receptor for SARS-CoV^12^ and COVID-19^13^. A groundbreaking research from University of Washington School of Medicine has proved that the expression of ACE2 increased as a function of viral load in COVID-19 patients^14^, which indicated that ACE2 might play a role in the disease progression and might become a target for therapy.

Before getting down to our own research, we noticed the statistical results of 989 hospitalized patients with COVID-19 pneumonia in Tongji Hospital (**Supplementary Table 1**). It was shown that the severity rate was linearly correlated with CFR and blood oxygen classification. There were significant differences between patients with Oximeter oxygen saturation (OOS) ⩽ 90 and patients with 90 < OOS ⩽ 94 in heart rate, eGFR and smoking history. Two vital cardiac biomarkers: cTnI, NT-proBNP were respectively up to 49.7% and 57.1% (*P*<0.001), while there were no differences in age, blood pressure and proportion of various underlying cardiovascular diseases. This phenomenon suggests that cardiac injury may be one of the important causes of death in patients with COVID-19 in the later stage of disease progression.

Another statistic demonstrates that the percentage of ACI during hospitalization was as high as 80% (Data on file) among all patients who died of disease. More importantly, 1/4 of the 619 patients with OOS>94 advanced to severe disease during hospitalization, and 28 (4.5% CFR) patients died, with cTnI, NT-proBNP exceeding 9.6% and 13.7%. We choose cTnI and NT-proBNP as the intermediate alternative indicators for ACI in 619 patients with OOS>94 and as prognostic factors as well. We hope the results can help to optimize the clinical pathway and treatment, and achieve the goal of strengthening early diagnosis and treatment to improve prognosis.

## Methods

### Study design and participants

619 COVID-19 inpatients admitted from January 27, 2020, to March 8, 2020 from Tongji Hospital, Tongji medical college of Huazhong University of Science and Technology (Wuhan, China) were included in this retrospective cohort. Considering the clinical significance, patients with basic heart failure or history of myocardial infarction were excluded. The diagnosis standard was based on Prevention and control Scheme for Novel Coronavirus Pneumonia (5^th^ edition) published by National Health Commission of the People’s Republic of China and WHO interim guidance. Current diagnostic tests for COVID-19 include RT-PCR assay of nasal and pharyngeal swab specimens for nucleic acid, positive serology for anti-COVID-19 specific IgM and/or IgG antibodies. For patients with the following symptoms: fever, sore throat, fatigue, coughing or dyspnea that are coupled with recent exposure, COVID-19 infection should be diagnosed with typical chest computerized tomography (CT) characteristics even though the RT-PCR results was negative^15^. Typical CT findings include bilateral pulmonary parenchymal ground-glass and consolidative pulmonary opacities, sometimes with a rounded morphology and peripheral lung distribution^16^. This study was on the basis of the principles of the Declaration of Helsinki.

### Data collection

Demographic data, medical history, clinical symptoms and signs, laboratory findings, chest CT, treatment, together with clinical results were obtained from electronic medical records. Clinical symptoms and signs consisted of fever, cough, dyspnea, myalgia, diarrhea, chest congestion, heart rate, blood pressure and OOS. Laboratory findings included a complete blood count, LDH, PT, APTT, Scr, eGFR, ALT, AST, serum albumin, D-dimer, NT-proBNP, TNI, CRP, ESR, IL-6, COVID-19 IgG, IgM and nucleic acid detection.

Drug use condition (anti-hypertension, antiviral drugs, antibiotics, corticosteroids, γ-globulin, traditional Chinese medicine (TCM)) were obtained from medical advice.

### Study definitions

ACRI was affirmed when cTnI level was >0.342 μg/L for males and >0.156 μg/L for females^17^ or NT-proBNP level was ⩾ 486 pg/mL. As the proportion of patients with two exceeded indicators increase continuously in patients with different blood oxygen grades (7.0%, 20.4%, 44.9%, **Supplementary Table 2**), we defined ACRI-stage-1 when only one of the two indicators is beyond the range. ACRI stage-2 was diagnosed when both of the two indicators were exceeded. The severity of COVID-19 was categorized as mild, common, severe, and critically ill according to *Diagnosis & Treatment Scheme for Novel Coronavirus Pneumonia (7*^*th*^ *edition)*. Oximeter oxygen saturation (OOS) is the fraction of oxygen-saturated hemoglobin relative to total hemoglobin in the blood. Saturation level that was ⩾94% or 90% - 94% or ⩽90% was respectively defined as OOS Level-I,II, III^18^.

### Statistical analysis

Continuous variables were presented as medians with standard deviation (SD) and compared with Student’s t test while categorical factors were expressed as frequency and counts and compared with Pearson χ^2^ test. Univariable and multivariable Cox regression models were applied to estimate odds ratio (OR) with 95% confidence interval (CI) to evaluate associations of demographic and clinical characteristics with COVID-19 in-hospital mortality. All analyses were conducted using SAS 9.4 (SAS Institute Inc., Cary, NC) or Stata 14 (Stata Corp, College Station, TX, US).

## Results

### Clinical characteristics of OOS Level- I COVID-19 patients categorized by ACRI stage

619 patients with OOS Level-I are classified based on acute cardiac related injury (ACRI) (**Table 1**). Three prognostic factors: the severity rate, CFR, LOS are in line with ACRI classification (**Figure 1**). There were 102 patients with ACRI stage-1 and 2, among which 25 patients died (CFR 24.5% for ACRI,15.2% for stage-1, 37.2%for stage-2, **Table 1, Figure 4**), 42 times of the CFR of patients without ACRI (0.58%). In terms of basic conditions and symptoms, we found that only SBP ⩾140 (44.2%, *P*=0.035) and dyspnea (32.5%, *P*=0.010) were associated with ACRI stage-2, while factors such as age, male, heart rate, ground glass shadows in both lungs and other major symptoms were not different. To our knowledge, this is the first clinical evidence that HSBP (not just HTN) and COVID-19 are related to disease progression. Other laboratory indicators were all linearly correlated with ACRI classification (**Supplementary Figure 1**). In the underlying disease characteristics, HTN, CHD, IKF (eGFR<60) are different statistically when DM and smoking history are distinct in ACRI stage-2. Patients receiving TCM treatment are scarce in ACRI stage-2 (*P*=0.001) but the portion of those treated with γ-globulin is relatively high in ACRI stage-1 (*P*=0.044) and stage-2 (*P*<0.001).

**Table 1.**
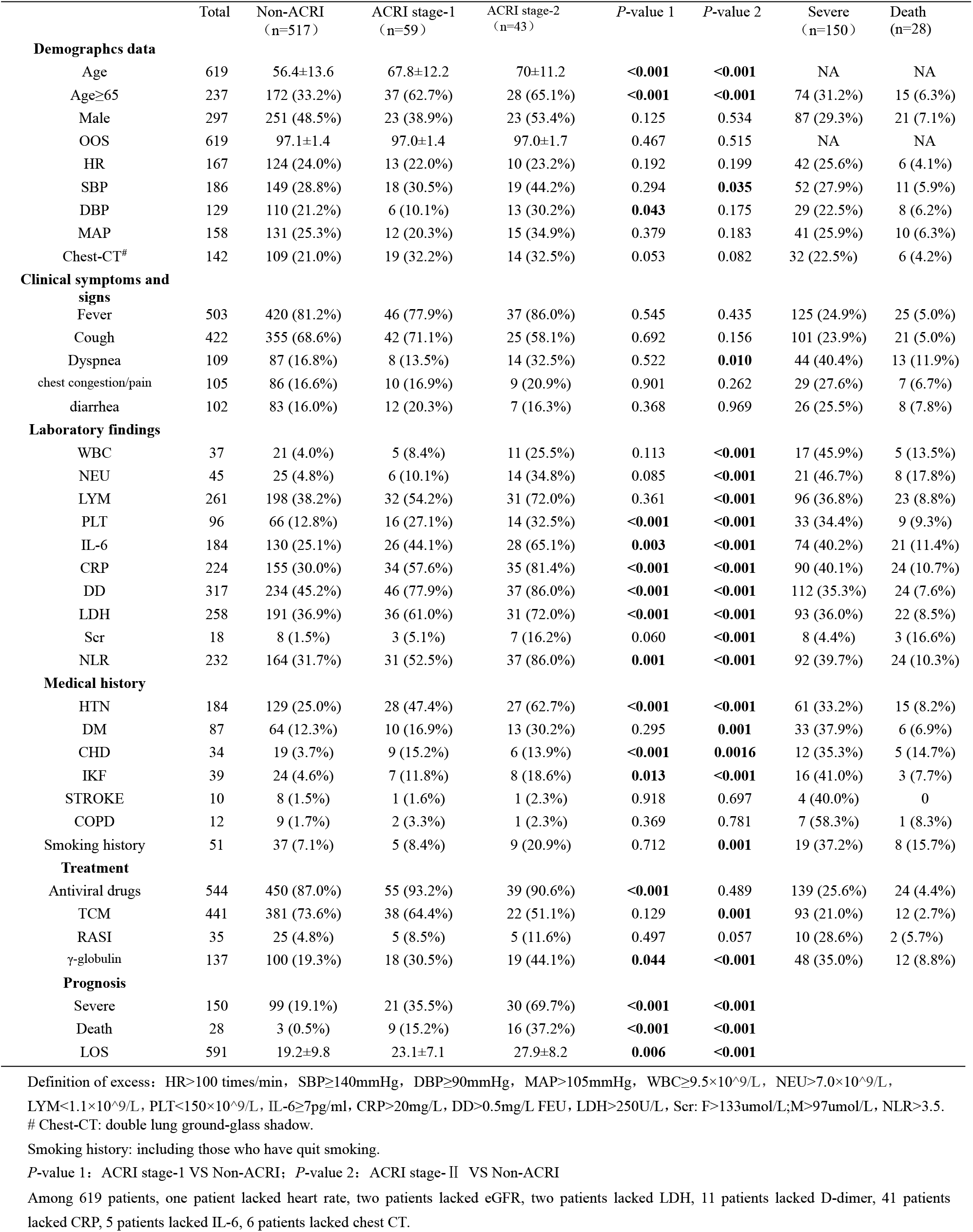
Clinical characteristics of patients with OOS Level-I (n=619)

|  | Total | Non-ACRI<br>(n=517) | ACRI stage-I<br>(n=59) | ACRI stage-2<br>(n=43) | P-value 1 | P-value 2 | Severe<br>(n=150) | Death<br>(n=28) |
| --- | --- | --- | --- | --- | --- | --- | --- | --- |
| **Demographics data** |  |  |  |  |  |  |  |  |
| Age | 619 | 56.4±13.6 | 67.8±12.2 | 70±11.2 | <0.001 | <0.001 | NA | NA |
| Age≥65 | 237 | 172 (33.2%) | 37 (62.7%) | 28 (65.1%) | <0.001 | <0.001 | 74 (31.2%) | 15 (6.3%) |
| Male | 297 | 251 (48.5%) | 23 (38.9%) | 23 (53.4%) | 0.125 | 0.534 | 87 (29.3%) | 21 (7.1%) |
| OOS | 619 | 97.1±1.4 | 97.0±1.4 | 97.0±1.7 | 0.467 | 0.515 | NA | NA |
| HR | 167 | 124 (24.0%) | 13 (22.0%) | 10 (23.2%) | 0.192 | 0.199 | 42 (25.6%) | 6 (4.1%) |
| SBP | 186 | 149 (28.8%) | 18 (30.5%) | 19 (44.2%) | 0.294 | 0.035 | 52 (27.9%) | 11 (5.9%) |
| DBP | 129 | 110 (21.2%) | 6 (10.1%) | 13 (30.2%) | 0.043 | 0.175 | 29 (22.5%) | 8 (6.2%) |
| MAP | 158 | 131 (25.3%) | 12 (20.3%) | 15 (34.9%) | 0.379 | 0.183 | 41 (25.9%) | 10 (6.3%) |
| Chest-CT<sup>#</sup> | 142 | 109 (21.0%) | 19 (32.2%) | 14 (32.5%) | 0.053 | 0.082 | 32 (22.5%) | 6 (4.2%) |
| **Clinical symptoms and signs** |  |  |  |  |  |  |  |  |
| Fever | 503 | 420 (81.2%) | 46 (77.9%) | 37 (86.0%) | 0.545 | 0.435 | 125 (24.9%) | 25 (5.0%) |
| Cough | 422 | 355 (68.6%) | 42 (71.1%) | 25 (58.1%) | 0.692 | 0.156 | 101 (23.9%) | 21 (5.0%) |
| Dyspnea | 109 | 87 (16.8%) | 8 (13.5%) | 14 (32.5%) | 0.522 | 0.010 | 44 (40.4%) | 13 (11.9%) |
| chest congestion/pain | 105 | 86 (16.6%) | 10 (16.9%) | 9 (20.9%) | 0.901 | 0.262 | 29 (27.6%) | 7 (6.7%) |
| diarrhea | 102 | 83 (16.0%) | 12 (20.3%) | 7 (16.3%) | 0.368 | 0.969 | 26 (25.5%) | 8 (7.8%) |
| **Laboratory findings** |  |  |  |  |  |  |  |  |
| WBC | 37 | 21 (4.0%) | 5 (8.4%) | 11 (25.5%) | 0.113 | <0.001 | 17 (45.9%) | 5 (13.5%) |
| NEU | 45 | 25 (4.8%) | 6 (10.1%) | 14 (34.8%) | 0.085 | <0.001 | 21 (46.7%) | 8 (17.8%) |
| LYM | 261 | 198 (38.2%) | 32 (54.2%) | 31 (72.0%) | 0.361 | <0.001 | 96 (36.8%) | 23 (8.8%) |
| PLT | 96 | 66 (12.8%) | 16 (27.1%) | 14 (32.5%) | <0.001 | <0.001 | 33 (34.4%) | 9 (9.3%) |
| IL-6 | 184 | 130 (25.1%) | 26 (44.1%) | 28 (65.1%) | 0.003 | <0.001 | 74 (40.2%) | 21 (11.4%) |
| CRP | 224 | 155 (30.0%) | 34 (57.6%) | 35 (81.4%) | <0.001 | <0.001 | 90 (40.1%) | 24 (10.7%) |
| DD | 317 | 234 (45.2%) | 46 (77.9%) | 37 (86.0%) | <0.001 | <0.001 | 112 (35.3%) | 24 (7.6%) |
| LDH | 258 | 191 (36.9%) | 36 (61.0%) | 31 (72.0%) | <0.001 | <0.001 | 93 (36.0%) | 22 (8.5%) |
| Scr | 18 | 8 (1.5%) | 3 (5.1%) | 7 (16.2%) | 0.060 | <0.001 | 8 (4.4%) | 3 (16.6%) |
| NLR | 232 | 164 (31.7%) | 31 (52.5%) | 37 (86.0%) | 0.001 | <0.001 | 92 (39.7%) | 24 (10.3%) |
| **Medical history** |  |  |  |  |  |  |  |  |
| HTN | 184 | 129 (25.0%) | 28 (47.4%) | 27 (62.7%) | <0.001 | <0.001 | 61 (33.2%) | 15 (8.2%) |
| DM | 87 | 64 (12.3%) | 10 (16.9%) | 13 (30.2%) | 0.295 | 0.001 | 33 (37.9%) | 6 (6.9%) |
| CHD | 34 | 19 (3.7%) | 9 (15.2%) | 6 (13.9%) | <0.001 | 0.0016 | 12 (35.3%) | 5 (14.7%) |
| IKF | 39 | 24 (4.6%) | 7 (11.8%) | 8 (18.6%) | 0.013 | <0.001 | 16 (41.0%) | 3 (7.7%) |
| STROKE | 10 | 8 (1.5%) | 1 (1.6%) | 1 (2.3%) | 0.918 | 0.697 | 4 (40.0%) | 0 |
| COPD | 12 | 9 (1.7%) | 2 (3.3%) | 1 (2.3%) | 0.369 | 0.781 | 7 (58.3%) | 1 (8.3%) |
| Smoking history | 51 | 37 (7.1%) | 5 (8.4%) | 9 (20.9%) | 0.712 | 0.001 | 19 (37.2%) | 8 (15.7%) |
| **Treatment** |  |  |  |  |  |  |  |  |
| Antiviral drugs | 544 | 450 (87.0%) | 55 (93.2%) | 39 (90.6%) | <0.001 | 0.489 | 139 (25.6%) | 24 (4.4%) |
| TCM | 441 | 381 (73.6%) | 38 (64.4%) | 22 (51.1%) | 0.129 | 0.001 | 93 (21.0%) | 12 (2.7%) |
| RASI | 35 | 25 (4.8%) | 5 (8.5%) | 5 (11.6%) | 0.497 | 0.057 | 10 (28.6%) | 2 (5.7%) |
| γ-globulin | 137 | 100 (19.3%) | 18 (30.5%) | 19 (44.1%) | 0.044 | <0.001 | 48 (35.0%) | 12 (8.8%) |
| **Prognosis** |  |  |  |  |  |  |  |  |
| Severe | 150 | 99 (19.1%) | 21 (35.5%) | 30 (69.7%) | <0.001 | <0.001 |  |  |
| Death | 28 | 3 (0.5%) | 9 (15.2%) | 16 (37.2%) | <0.001 | <0.001 |  |  |
| LOS | 591 | 19.2±9.8 | 23.1±7.1 | 27.9±8.2 | 0.006 | <0.001 |  |  |

Definition of excess: HR>100 times/min, SBP≥140mmHg, DBP≥90mmHg, MAP>105mmHg, WBC≥9.5×10<sup>9</sup>/L, NEU>7.0×10<sup>9</sup>/L, LYM<1.1×10<sup>9</sup>/L, PLT<150×10<sup>9</sup>/L, IL-6≥7pg/ml, CRP>20mg/L, DD>0.5mg/L FEU, LDH>250U/L, Scr: F>133umol/L;M>97umol/L, NLR>3.5. <sup>#</sup> Chest-CT: double lung ground-glass shadow.
Smoking history: including those who have quit smoking.
P-value 1: ACRI stage-I VS Non-ACRI; P-value 2: ACRI stage- II VS Non-ACRI
Among 619 patients, one patient lacked heart rate, two patients lacked eGFR, two patients lacked LDH, 11 patients lacked D-dimer, 41 patients lacked CRP, 5 patients lacked IL-6, 6 patients lacked chest CT.

### Analysis of risk factors associated with ACRI in OOS Level-I COVID-19 patients

In order to further reveal correlation intensity between ACRI and the factors above, we selected SBP, dyspnea, and other laboratory indexes as indicators for hospitalization, including three indexes reflecting basic conditions: age, underlying diseases, medicines use for treatment. Based on two distinct patterns (1, ACRI stage-1 + stage-2 as dependent variable; 2, ACRI stage-2 as dependent variable), we conduct multivariate regression statistics respectively.

**Figure 1.**
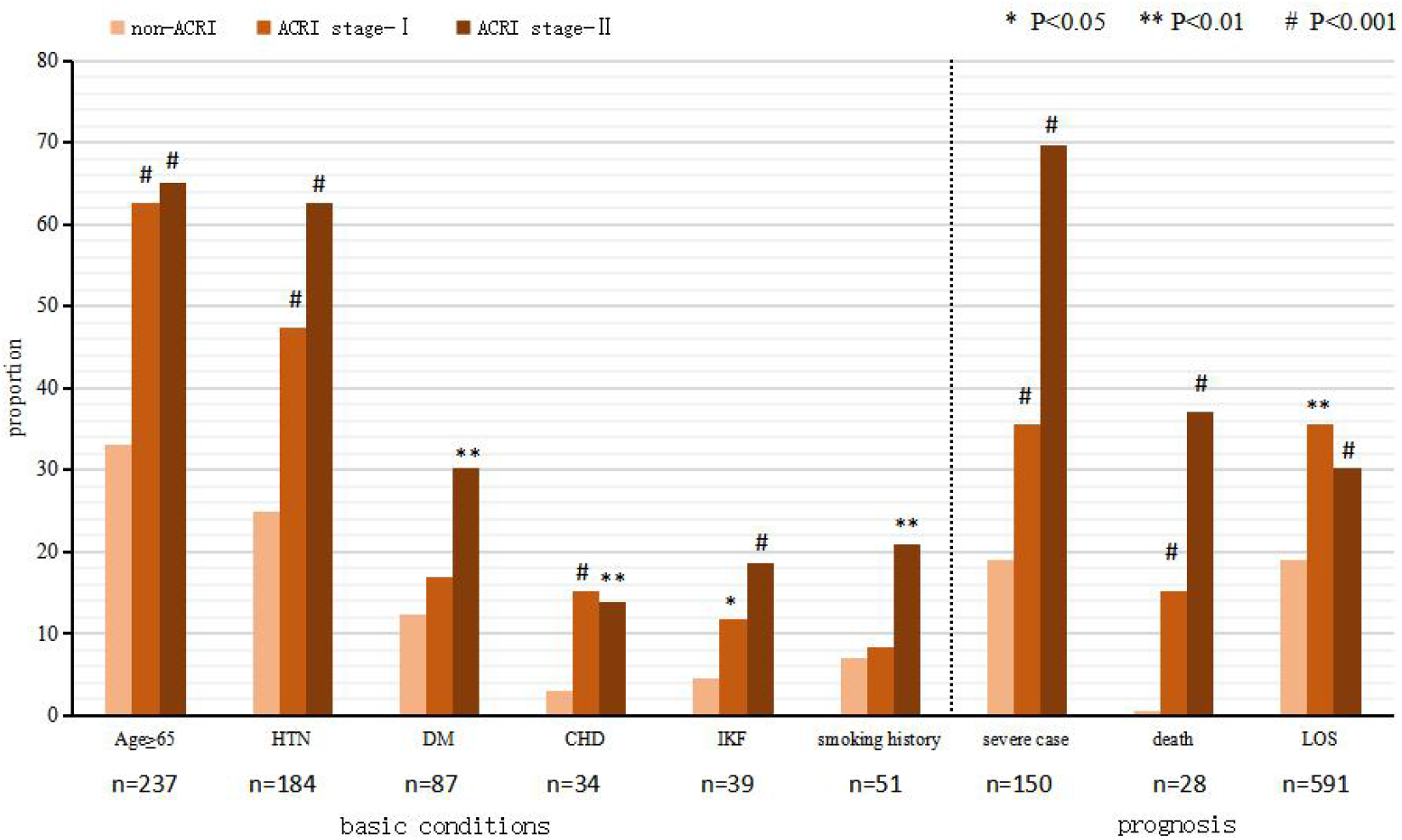
The comparison of basic conditions, prognostic factors and heart injury among OOS Level-I patients (n=619)

The results illustrated that only PLT < 150 and NLR > 3.5 show significant differences among 12 single-factor-related indicators (**Figure 2**). Among 9 single factor – related or clinically meaningful indicators for basic conditions, four indexes: elderly, HTN, TCM andγ-globulin treatment are significantly different in both patterns (**Figure 2**), with their adjusted OR with ACRI (including stage 1 and 2) respectively at 2.83(*P*<0.001),2.09(*P*=0.005), 0.55(*P*=0.017),2.08(*P*=0.004). CHD is only correlated to ACRI while DM and smoking history are only related to ACRI stage-2 (**Figure 2**). Distinct from the cohort study introduced in background, the percentage of patients with IKF (eGFR< 60) in pattern-1 has no significantly difference from that in pattern-2 (S**upplementary Figure 1**), which suggests that the pathological mechanism of early ACRI in COVID-19 patients might be independent from the kidney-RAS system.

**Figure 2.**
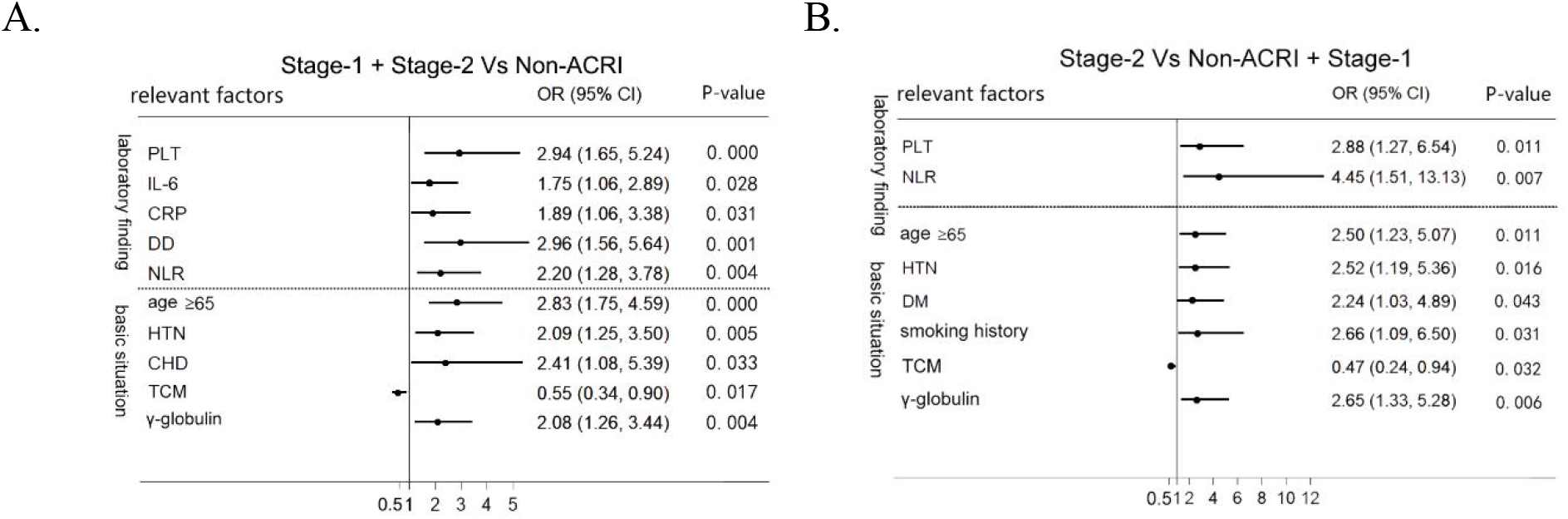
Multivariable Cox regression of risk factors for ACRI. (Shows risk factors with *P* < 0.05 only). (A) In multivariable pattern-1: ACRI stage-1+stage-2 (n=102) VS Non-ACRI (n=517) (B) In multivariable pattern-2: ACRI stage-2 (n=43) VS Non-ACRI+Stage-1 (n=576)

In 102 patients with ACRI, we observed those whose PLT >150 or <150, focusing on related indexes like heart rate,SBP,CRP,LDH,D-dimer,eGFR (**Supplementary Table 3**).No significant difference was found between the two groups, further verifying that the decrease of PLT shown in multiple regression statistics might be an independent risk factor of ACRI.

### Subgroup analysis of related factors affecting the outcomes of patients with early ACRI (N =102)

From the subgroup analysis of related factors and three prognostic indicators in 102 ACRI patients, we found that ACRI occurred in 102 of 619 patients admitted into hospital, 51 patients (50%) developed into severe disease, and nearly 1/4 patients with ACRI died (89% of total number of death).

Based on the results above, we selected elderly, SBP, PLT, NLR, γ-globulin as ACRI related factors and add another six possible indexes into analysis: TCM, male, heart rate, chest distress and/or chest pain, eGFR, corticosteroids therapy, RASI treatment. The results (**Table 2**) show that, in LOS indexes, only γ-globulin treated group (29.0 ± 7.2 vs. 23.5 ± 8.1 days (*P* = 0.004)) and NLR >3.5 group (27.1 ± 7.7 vs. 22.5 ± 8.1 days (*P* = 0.013)) show differences. Besides, there were statistical differences(*P* < 0.005)in the proportion of severe and/or death patients among the following groups (**Figure 3**): male, SBP ⩾ 160, NLR>3.5, corticosteroids therapy, TCM. However, elderly, heart rate, chest distress/chest pain, PLT<150, eGFR, RASI treatment had no difference in three prognostic indicators: the severity rate, CFR, LOS (*P* >0.05). Moreover, the mortality of non-elderly patients was 32.4%, higher than the elderly (20%), which could partly explain the phenomenon that middle-aged men were reported to have a higher mortality in some American countries^19, 20^.

**Table 2.** Indicators of prognosis for ACRI patients with OOS Level-I (n=102)

| Factors | Total | Severe (n=51) | Death (n=25) | LOS (n=77) |
| --- | --- | --- | --- | --- |
| **Age** |  |  |  |  |
| <65 | 37 | 20 (54.0%) | 12 (32.4%) | 24.6±9.7 |
| ≥65 | 65 | 31 (47.9%) | 13 (20.0%)<sup>#</sup> | 25.8±7.5 |
| **Gender** |  |  |  |  |
| female | 56 | 21 (37.5%) | 6 (10.7%) | 24.6±8.4 |
| male | 46 | 30 (65.2%)<sup>A</sup> | 19 (41.3%)<sup>A</sup> | 27.0±7.8 |
| **HR** |  |  |  |  |
| <100 | 78 | 39 (50.0%) | 20 (25.6%) | 24.9±8.6 |
| ≥100 | 24 | 12 (50.0%) | 5 (20.8%) | 26.9±6.9 |
| **SBP pattern-1** |  |  |  |  |
| <120 | 22 | 11 (50.0%) | 6 (27.2%) | 25.5±8.0 |
| 120-139 | 43 | 18 (41.8%) | 9 (20.9%) | 24.5±8.7 |
| 140-159 | 24 | 10 (41.6%) | 6 (25.0%) | 24.8±7.0 |
| ≥160 | 13 | 12 (92.3%) | 4 (30.7%) | 29.5±7.8 |
| **SBP pattern-2** |  |  |  |  |
| <160 | 89 | 39 (43.8%) | 21 (23.5%) | 24.9±8.2 |
| ≥160 | 13 | 12 (92.3%)<sup>B</sup> | 4 (30.7%)<sup>B</sup> | 29.5±7.8<sup>B</sup> |
| **chest congestion/pain** |  |  |  |  |
| No | 83 | 40 (48.1%) | 19 (22.8%) | 25.5±8.0 |
| Yes | 19 | 11 (57.8%) | 6 (31.5%) | 25.0±9.1 |
| **PLT** |  |  |  |  |
| ≥150 | 72 | 33 (45.8%) | 17 (23.6%) | 24.7±8.0 |
| <150 | 30 | 18 (60.0%) | 8 (26.6%) | 27.1±8.7 |
| **NLR** |  |  |  |  |
| ≤3.5 | 34 | 11 (32.3%) | 4 (11%) | 22.5±8.1 |
| >3.5 | 68 | 40 (58.8%)<sup>C</sup> | 21 (30.8%)<sup>C</sup> | 27.1±7.7<sup>C</sup> |
| **eGFR** |  |  |  |  |
| ≥90 | 30 | 19 (63.3%) | 11 (36.6%) | 25.9±7.9 |
| 60-90 | 57 | 23 (40.3%) | 11 (19.2%) | 24.8±8.4 |
| <60 | 15 | 9 (60.0%) | 3 (20.0%) | 26.8±8.4 |
| **γ-globulin** |  |  |  |  |
| No | 65 | 30 (46.1%) | 15 (23.1%) | 23.5±8.1 |
| Yes | 37 | 21 (56.8%) | 10 (27.0%) | 29.0±7.2<sup>D</sup> |
| **corticosteroids** |  |  |  |  |
| No | 53 | 20 (37.7%) | 6 (11.3%) | 24.1±8.1 |
| Yes | 49 | 31 (63.3%)<sup>E</sup> | 19 (38.8%)<sup>E</sup> | 27.5±8.0 |
| **RASI** |  |  |  |  |
| No | 92 | 45 (48.9%) | 23 (25.0%) | 25.5±8.4 |
| Yes | 10 | 6 (60.0%) | 2 (20.0%) | 24.8±6.6 |
| **TCM** |  |  |  |  |
| No | 42 | 29 (69.0%) | 14 (33.3%) | 24.6±8.1 |
| Yes | 60 | 22 (36.7%)<sup>F</sup> | 11 (18.3%)<sup>F</sup> | 27.0±8.4 |

P-value (Yes Vs No): A. Severe $P=0.005$ Death $P<0.001$
B. Severe $P<0.001$ Death $P=0.579$ LOS $P=0.113$
C. Severe $P=0.005$ Death $P=0.007$ LOS $P=0.013$
D. LOS $P=0.004$
E. Severe $P=0.010$ Death $P=0.001$
F. Severe $P=0.027$ Death $P=0.084$

**Figure 3.**
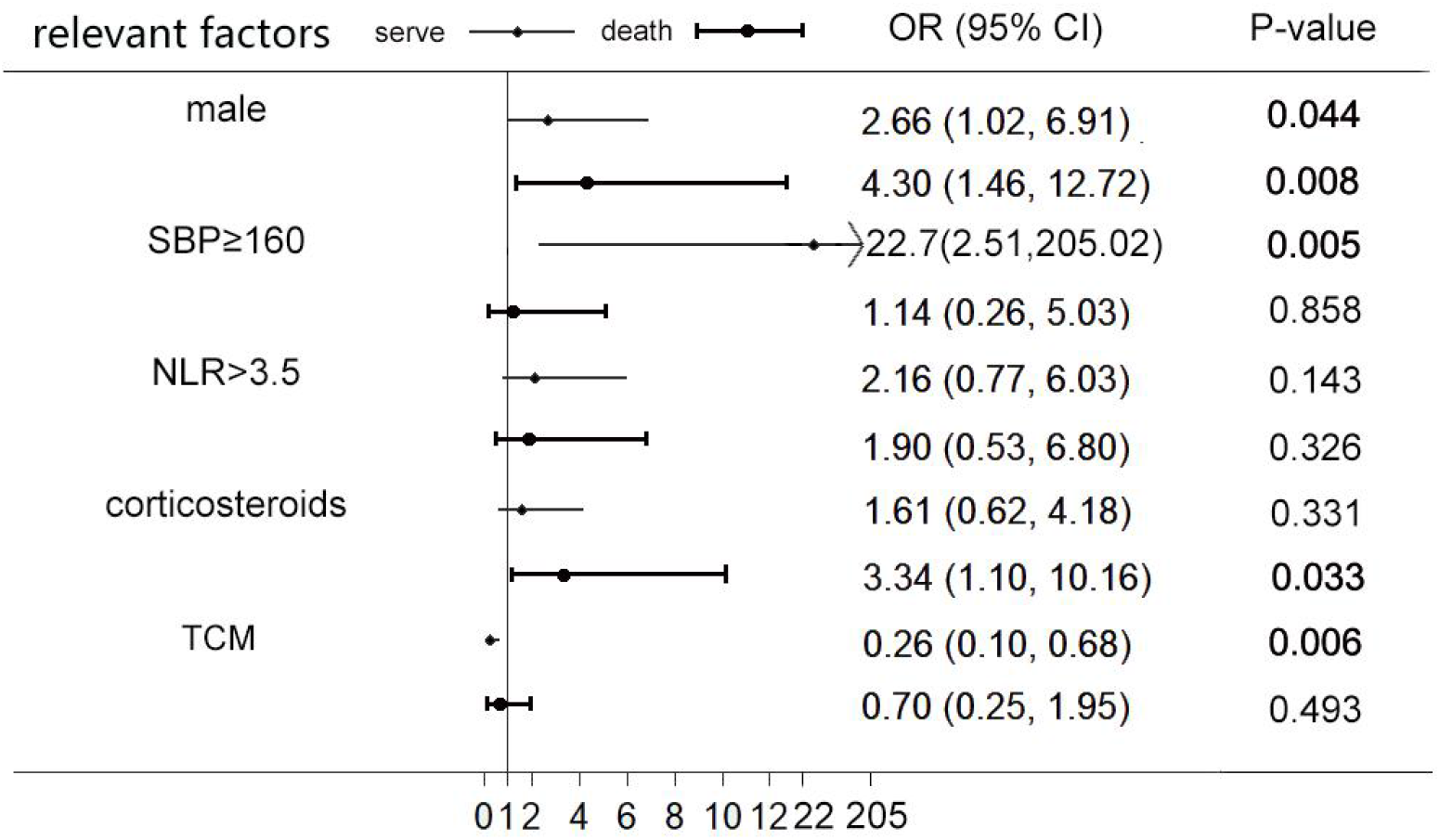
Multivariable regression of risk factors for severe and dead ACRI patients (n=102)

### Note

Although the antiviral treatment was associated with ACRI stage-1, considering that most patients in this group had been treated with this therapy after admission and the type and dosage of antiviral medicine vary greatly, we did not think it was sufficient to suggest that the antiviral treatment was associated with ACRI, thus it was not introduced into multiple regression statistics.

Given the clinical emphasis on whether RASI has an additional cardiac protective effect, no difference was found between the two models for RASI treatment, whether single-factor or multivariate, but regression statistics were retained and forced into the data.

## Discussion

A presentation on the current worldwide pandemic caused by COVID-19 was given by Anthony Fanci, an expert from American National Institute of Allergy and Infectious Diseases on January 23, 2020. The presentation listed risk factors for severe COVID-19 illness, and the top three were age above 65 years, living in nursing homes or long-term care facilities and hypertension. Also, the top three manifestations of severe COVID-19 disease include ARDS, hyper-inflammation and acute cardiac injury, arrhythmias, cardiomyopathy, which suggests that cardiovascular system injury may play an important role in COVID-19 severe patients^21^. Furthermore, a meta-analysis including 1664 patients illustrated that ACI could occur in one third of patients with COVID-19^22^. More than one studies have demonstrated that Cardiac injury was independently associated with significantly increase of mortality^22, 23^. Considering the above conclusion, we believe early diagnosis and treatment will be much important to improve the prognosis of ACI patients with COVID-19. Therefore, we selected patients with OOS-level I (not using a ventilator) to analyze, expecting to find risk factors of ACRI at early stage, monitor factors associated with disease progression and provide evidence for early clinical treatment.

### Occurrence of ACRI in COVID-19

This retrospective study provided comprehensive data on the basic conditions, clinical symptoms, laboratory examinations and outcomes of 619 hospitalized COVID-19 patients in Tongji hospital. A recent meta-review revealed that the OR of patients with ACI and finally died was 3.41. The risk rate varies from 1.30 to 11.40 based on several researches^22^ and our study shows the number of ACRI patients who died was 42 times that of those without ACRI (**Table 1**).

This study revealed the difference between SARS-CoV-2 and SARS. Study has shown that SARS-CoV-2 has a higher binding strength for ACE2 than SARS ^24^, but most of the infected people were clinically asymptomatic^25^, which means many infected people will not develop organ damage, while symptomatic patients had longer course of disease than SARS^26^. We conjectured that higher binding strength extended the distribution of ACE2 in the host, changing the time-dynamic characteristics of the virus causing pathological changes in different target organs, affecting the length of virus replication and the virus tolerance of individual patients, resulting in some non-severe patients underlying basic cardiovascular risks suffered from heart injury at admission.

### Risk factors for ACRI and disease progression

The exact mechanism of ACRI in COVID-19 is unclear, and multiple factors may be involved such as direct viral invasion, severe hypoxia, poor perfusion, formation of microthrombi in the coronary arteries and cytokine storm^27^.

Multivariate regression analysis in two patterns illustrated that only PLT<150 (*P*=0.011, OR=2.882) and NLR>3.5 (*P*=0.007, OR=4.448) were related to ACRI, while other inflammation indicators are not independent. To our knowledge, this study is the first one specialized the association between reduction of platelet and COVID-19 aggravation, while the mechanism of platelet involved in the pathogenesis of ACRI in COVID-19 patients is unclear. A previous study from Jinyingtan Hospital, Wuhan, China, containing 1476 consecutive patients, had reported that PLT reduction was associated with an increased risk of in-hospital mortality in patients with COVID-19^28^, while in our study, PLT<150 is not associated with mortality. The results indicated PLT reduction might be a sign of lung injury^29^ since a previous study published on *Nature* provided direct evidence that the lung was a major site of platelet biogenesis and produced approximately 50% of total platelet^30^. We supposed the platelet may be served as a bridge connecting lung injury and cardiac injury. Platelets act as immunomodulatory cells that contact their role via numerous mechanisms^31^. On the one hand, the inflammatory storms caused by COVID-19 may contribute to lung injury and heart injury through platelet-leukocyte-interaction. The lungs are most vulnerable organs to inflammatory storms because of the extensive capillary bed and abundant immune cells. Pro-inflammatory factors act as a chemotactic signal to recruit circulating leukocytes, which accumulate inappropriately in lung tissue^32^. During the processes, platelets may be activated, lead adhesive protein to bind leukocytes and exert strong pro-inflammatory effects that damage tissue. It can be corroborated by the depleting platelets in animal models of lung injury ameliorating tissue damage^33^. As for the heart injury, it has been known that the interaction between platelets and leukocytes is associated with MI^34^. Neumann et al. found that leukocyte platelet adhesion was increased in the peripheral venous blood samples of patients with AMI, and this was part of the regulation of the inflammatory response in AMI ^35^. In our study, the percentage of patients with high WBC and NEU in ACRI stage-2 was significantly higher than those in non-ACRI, which also indicated that platelets-leukocytes-interaction may connect lung injury with heart injury and play a role in the ACRI patients. One the other hand, despite the negative effects of platelet-leukocyte-interaction, platelets can release some constituents that may have beneficial effects on the integrity of the coronary endothelium and on cardiac function^36, 37^. The cardioprotective constituents include serotonin, thromboxane A2, sphingosine-1-phosphate (S1P), the platelet α granule contents transforming growth factor-beta1 (TGF-β1) and so on^38, 39^. In summary, the lung injury caused by COVID-19 results in the reduction of platelet synthesis, and the reduction of platelet results in the downregulation of these cardioprotective molecules, which may aggravate myocardial damage.

NLR (*P*=0.007, OR=4.448) is independently related to ACRI. Neutrophil (NEU) is a major component of the leukocyte population, which can activate and migrate from the venous system to the immune organ or system. On the one hand, virus-related inflammatory factors, such as IL-6, IL-8, TNF-α, GCSF and IFN-γ can trigger NEU. On the other hand, viral infection triggering immune response mainly relies on lymphocyte^40^, which significantly decreases CD4^+^ T lymphocytes and increases CD8^+^ suppressor T lymphocyte. This mechanism can explain why NLR elevated in COVID-19. Prognosis risk factor analysis indicated that immune mechanism had no special effect on prognosis of ACRI, prompting that the recognized “inflammatory storm” might not be the main reason for the worse prognosis of ACRI patients. The patients might die from cardiac-RAS imbalance rather than heart damage, and the exact mechanism still needs to be further clarified.

Smoking history was also found to be associated with ACRI stage-2 (*P*<0.001, OR=2.94). To the best of our knowledge, it is the first time that smoking history has been associated with increasing risk of acute cardiac injury.

Combining the results of risk factors for ACRI and prognosis, we found that HSBP played a role in the disease progression. Our results demonstrated SBP⩾140 (*P*=0.035, ACRI stage-2 VS non-ACRI, OR=1.53) was associated with ACRI stage-2 and SBP ⩾ 160 (*P*=0.005, OR=22.70) is independent risk factor for severe disease. To our knowledge, this is the first clinical evidence that HSBP (not just HTN) are related to COVID-19 disease progression. Previous study had found the prevalence of hypertension in severe patients was significantly higher than in non-severe patients^41^. We supposed that vascular damage may exist in the HSBP patients, the vascular damage directly leaded to ACRI, which made HSBP a risk factor for ACRI. One possible explanation is the impaired endothelial function and ACE2. ACE2, a functional receptor for SARS-CoV and COVID-19, was reported to be a protective factor against SARS-CoV-induced lung injury^42, 43^. Previous study found that ACE2 was prominently expressed by cardiac pericytes in human heart and airway epithelia cells in the lung ^44^. The respiratory epithelium is the most common entry point for viral infection. The virus enters the blood circulation from the tissue in reverse. The pericytes are normally shielded from the blood behind an intact endothelial barrier, impaired endothelial barrier function caused by hypertension or DM allowing SARS-CoV-2 to reach and infect the pericytes^44^. Immune-attack of the infected pericytes lead to microvascular inflammation, which exacerbates vascular leakage and endothelial pro-inflammatory and pro-thrombotic response^45^. The following thrombogenesis and thromboembolism in the coronary may contribute to ACRI.

Another explanation for the relationship between HSBP and ACRI is RAS imbalance. A previous research which monitored the dynamic change of blood pressure every day of COVID-19 patients revealed that severe cases had higher level of SBP and that the prevalence rate of new onset hypertension was significantly higher in hospitalized severe patients with COVID-19^46^. These results suggested that new onset high blood pressure and ACRI might be caused by RAS imbalance, while RAS imbalance might be mediated by ACE2. We suppose that COVID-19 and HTN promote each other and aggravate lung and heart injury. This fact may partly explain the new onset hypertension in severe COVID-19 patients. Whether it is vascular damage or RAS imbalance that relates the HBP and ACRI, it is far more different from the recognized inflammatory factors.

The multivariate regression statistics of prognostic risk factors demonstrated that CHD had no effect on the prognosis, which is consistent with a previous study from Metropolitan Atlanta, Georgia, March–April 2020^47^. The results indicated that basal peripheral vascular function is more relevant to ACRI in COVID-19 patients than coronary disease. Furthermore, IKF (eGFR) was not associated with ACRI at early stage of the progression, while senior age, smoking and HSBP associated. These results were totally different from a previous study from Renmin Hospital of Wuhan University containing 671 hospitalized patients that demonstrated senior age and comorbidities including hypertension, coronary heart disease, chronic renal failure, and chronic obstructive pulmonary disease were all predictors of myocardial injury^7^. It can be considered that high-risk cardiovascular patients with factors related to vascular damage (including senior age, smoking, and HSBP) are equally or more dangerous than patients with cardiovascular target organ damage history (including CHD, IKF and stroke) in the early stage of the disease, which is contrary to the traditional cardiovascular risk chain, and this may also be the reason for the rapid deterioration of many patients who were not previously considered dangerous. In summary, we speculate that ACRI is not only a “suspected heart injury", but also an early sign of “multi-organ injury” and “vascular function injury".

Male was also found to be related with severe cases and death, which can be explained by the lack of estrogen. A research from Otto-von-Guericke University Magdeburg has proved estrogen could increase the expression of ACE2 on the blood vessel and achieve protective effects in atrial myocardium^48^. Estrogen deficiency means high vascular risk, which may contribute to ACRI.

### Effects of medical treatment

A meaningful research from University of Washington School of Medicine proved that the expression of ACE2 increased as a function of viral load in COVID-19 patients^14^. The conclusion may explain why some non-cardiovascular medicine treatments may affect the development of the disease by changing the expression of ACE2.

γ-globulin treated group showed difference in LOS indexes (29.0 ± 7.2 days Vs 23.5 ± 8.1 days, *P*=0.004) and associated with ACRI stage-2 (*P*=0.006, OR=2.651). The result was opposite to a report which included 58 severe cases and demonstrated IVIG could improve the patients’ indicators and the treatment efficiency ^49^. A previous study found that in wild-type mice, exposure to high-protein diet was associated with an increase in renal blood flow, renal mass, renal expression of (pro)renin and intrarenal ACE2 activity, while in ACE2 KO mice, high-protein diet caused a functional loss of the renal ^50^. We believe that γ-globulin treatment may lead to ACRI and prolong the detox time by affecting ACE2 expression in target organ. The two findings are firstly reported to explain why γ-globulin should not be used for treatment in Covid-19 patients with high cardiovascular risk, and the preventive recommendation of increasing high-protein intake to improve inflammatory resistance may not be suitable for all people.

TCM treatment is protective factor for severe cases, but it had no effect on LOS and death. It was suggested that its positive effect may not be directly related to the suppression of viruses, but avoided the side effects of NASID and improved symptoms^51^. RASI treatment (OR=1.446, *P*=0.401, **Supplementary Table 4**) brought no favorable results in early stage. It was different from the results of a previous study^16^ that inpatient use of ACEI/ARB was associated with lower risk of all-cause mortality compared with ACEI/ARB nonusers. The difference can be explained by the different enrollment of patients.

Corticosteroids treatment (*P*=0.033, OR=3.34) was associated with increased mortality. This was consistent with a recently published study from the University of Oxford’s RECOVERY trial that corticosteroids increasing the mortality of non-oxygenated hospitalized patients^52^, suggesting that an empirical protocol based on the severity of the illness does not ensure patient benefit. The ROCOVERY study has shown that ventilator-treatment can reduce 28-day in-hospital mortality^52^, but it is unclear whether specific patients with heart injury will benefit.

As for antithrombotic treatment, although National Health Commission of the People’s Republic of China recommended heparin use in severe patients^53^, most patients enrolled in this study were in the early stage of the epidemic, so no observational analysis was performed. We found that PLT reduction had no effect on the ACRI prognosis, including severe rate, mortality and LOS. This result is consistent with a recent British study about brain damage (only 9 of 57 ischemic stroke hemorrhagic strokes)^54^. This is good news, indicating that the common increase in DD and decrease in PLT in Covid-19 hospitalized patients may not be a systemic coagulation dysfunction in the traditional sense, and most patients can improve prognosis by strengthening anticoagulation therapy.

According to the strategy of ‘early diagnosis and early treatment’ proposed by WHO^55^, the non-severe patients in our cohort have achieved early treatment in hospital. Despite some patients (70/517, 13.5%) had to suffer longer LOS (>30 days), the survival rate of non-ACRI patients reached very high level (99.4%, **Figure 4**). On the contrary, the mortality of patients with ACRI was still unsatisfactory (24.5%), so we need to focus on high-risk factors identification. Early diagnosis and treatment may optimize the clinical pathway and guidance.

**Figure 4.**
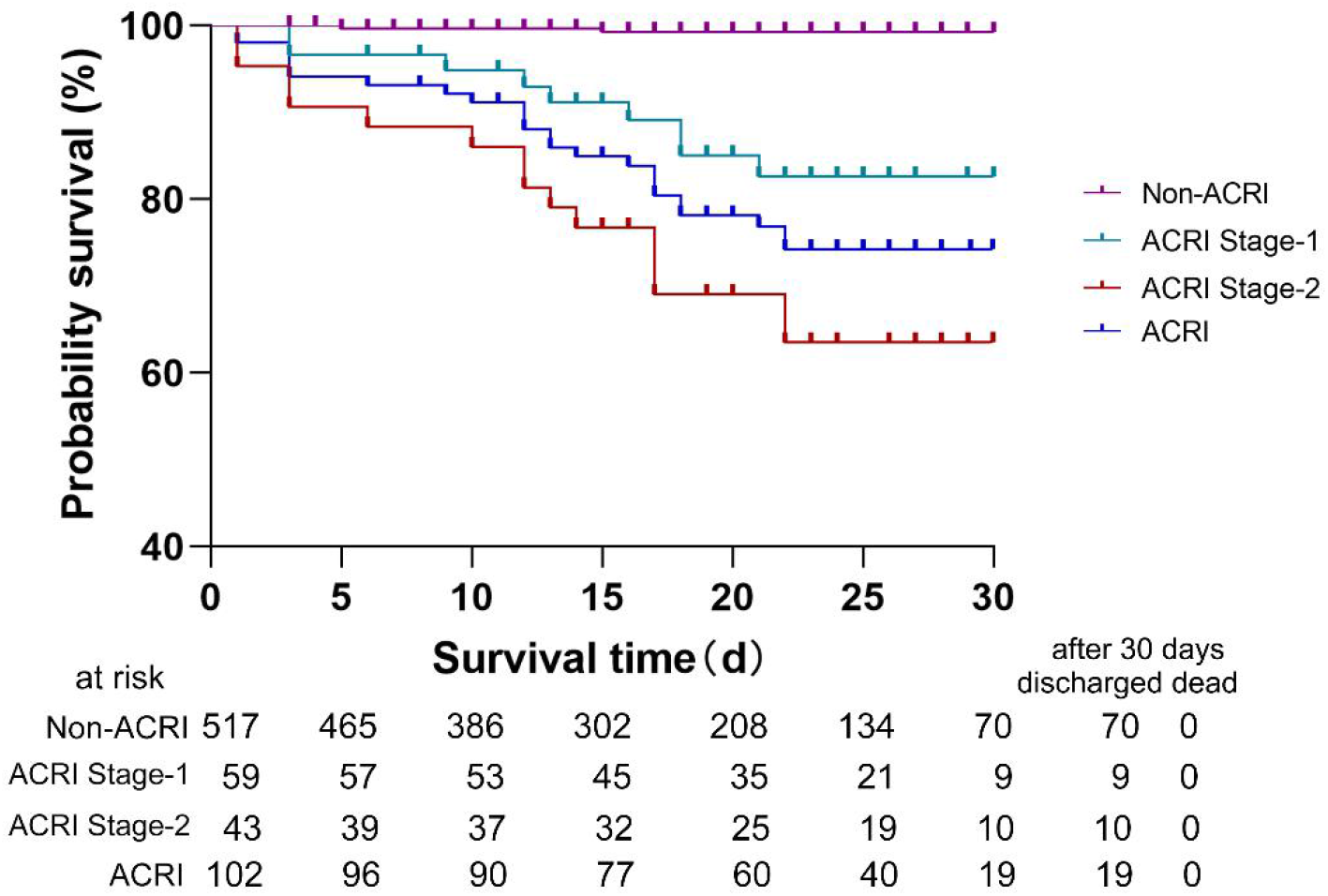
The probability of survival in 30 days among patients with Non-ACRI, ACRI Stage-1, ACRI Stage-2 and ACRI

### Limitations

This study has some limitations. First, due to the retrospective study design, not all laboratory tests were done in all patients, and the laboratory index testing for each patient was measured at different time. Second, due to special circumstances during the outbreak, arterial blood gas test, electrocardiography and echocardiography data were lacking for many cases, which limits the assessment of the extent of ACRI. Third, we did not monitor the dynamic change of blood pressure, which made this study lack of evidence of new onset hypertension to support our hypothesis. Last, it was a single-center study. The number of samples is not big enough and selection bias may exist, thus further prospective multi-centric clinical studies are warranted to further confirm the findings obtained in the present study.

## Data Availability

The raw data was extracted from electronic medical records with standardized data collection forms in Tongji Hospital (Wuhan, China), which had been approved by the Research Ethics Commission of Tongji Hospital, Tongji medical college of HUST. The data used to support the findings of this study are available from the corresponding author upon request.

## Nonstandard Abbreviations and Acronyms

ACI: Acute cardiac injury
ACRI: Acute cardiac related injury
AMI: Acute myocardial injury
CFR: Case-fatality rate
CHD: Coronary heart disease
COVID-19: Coronavirus disease-2019
CRP: C-reactive protein
cTnI: Cardiac troponin I
CVD: Cardiovascular disease
DD: D-Dimer
DM: Diabetes mellitus
eGFR: Estimated glomerular filtration rate
HSBP: High systolic blood pressure
IKF: Impaired kidney function
IL-6: Interleukin 6
IVIG: Intravenous immunoglobulin
LDH: Lactate dehydrogenase
LOS: Length of stay
MAP: Mean arterial pressure
NEU: Neutrophil
NT-proBNP: N-terminal pro-brain natriuretic peptide
NLR: Neutrophil lymphocyte ratio
OOS: Oximeter oxygen saturation
RAS: Renin-angiotensin-aldosterone system
RASI: Renin-angiotensin-aldosterone system inhibitors
Scr: Serum creatinine
WHO: World Health Organization

## Acknowledgements

We thank Professor Weizhong Zhang from Shanghai Institute of Hypertension,Ruijin Hospital, Shanghai Jiaotong University, Shanghai, China for support. We are also grateful for all COVID-19 patients for their participation in our research.

## Authors’ Contributors

Hesong Zeng, Yin shen conceived the study and its design, had full access to the data, and take responsibility for the integrity of the data and accuracy of the analysis. Xueli Wang, Baige Li and Tianlu Zhang contributed to data analyses. Sichun Chen and Yang Xie drafted the manuscript. All authors approved the final version of the article.

